# Renin-angiotensin system blockers and susceptibility to COVID-19: a multinational open science cohort study

**DOI:** 10.1101/2020.06.11.20125849

**Authors:** Daniel R. Morales, Mitchell M. Conover, Seng Chan You, Nicole Pratt, Kristin Kostka, Talita Duarte-Salles, Sergio Fernández-Bertolín, Maria Aragón, Scott L. DuVall, Kristine Lynch, Thomas Falconer, Kees van Bochove, Cynthia Sung, Michael E. Matheny, Christophe G. Lambert, Fredrik Nyberg, Thamir M. Alshammari, Andrew E. Williams, Rae Woong Park, James Weaver, Anthony G. Sena, Martijn J. Schuemie, Peter R. Rijnbeek, Ross D. Williams, Jennifer C.E. Lane, Albert Prats-Uribe, Lin Zhang, Carlos Areia, Harlan M. Krumholz, Daniel Prieto-Alhambra, Patrick B. Ryan, George Hripcsak, Marc A. Suchard

## Abstract

**Introduction:** Angiotensin converting enzyme inhibitors (ACEs) and angiotensin receptor blockers (ARBs) could influence infection risk of coronavirus disease (COVID-19). Observational studies to date lack pre-specification, transparency, rigorous ascertainment adjustment and international generalizability, with contradictory results.

**Methods:** Using electronic health records from Spain (SIDIAP) and the United States (Columbia University Irving Medical Center and Department of Veterans Affairs), we conducted a systematic cohort study with prevalent ACE, ARB, calcium channel blocker (CCB) and thiazide diuretic (THZ) users to determine relative risk of COVID-19 diagnosis and related hospitalization outcomes. The study addressed confounding through large-scale propensity score adjustment and negative control experiments.

**Results:** Following over 1.1 million antihypertensive users identified between November 2019 and January 2020, we observed no significant difference in relative COVID-19 diagnosis risk comparing ACE/ARB vs CCB/THZ monotherapy (hazard ratio: 0.98; 95% CI 0.84 - 1.14), nor any difference for mono/combination use (1.01; 0.90 - 1.15). ACE alone and ARB alone similarly showed no relative risk difference when compared to CCB/THZ monotherapy or mono/combination use. Directly comparing ACE vs. ARB demonstrated a moderately lower risk with ACE, non-significant for monotherapy (0.85; 0.69 - 1.05) and marginally significant for mono/combination users (0.88; 0.79 - 0.99). We observed, however, no significant difference between drug-classes for COVID-19 hospitalization or pneumonia risk across all comparisons.

**Conclusion:** There is no clinically significant increased risk of COVID-19 diagnosis or hospitalization with ACE or ARB use. Users should not discontinue or change their treatment to avoid COVID-19.

## Introduction

People with cardiovascular diseases and hypertension are more likely to develop severe complications of coronavirus disease 2019 (COVID-19) resulting in hospitalization and death (Shi et al. 2020; Guo et al. 2020); (Ruan et al. 2020). Speculatively, angiotensin converting enzyme inhibitors (ACEs) and angiotensin-II receptor blockers (ARBs), both renin-angiotensin system (RAS) blockers, may influence susceptibility to COVID-19 and worsen its severity. Driving this hypothesis is the mechanism that SARS-CoV-2 enters human cells by binding to the membrane-bound aminopeptidase angiotensin-converting enzyme 2 (ACE2) (Li et al. 2003; Hoffmann et al. 2020), for which chronic exposure to RAS therapy may alter expression (Ferrario et al. 2005; Vuille-dit-Bille et al. 2015; Soler et al. 2009; Sukumaran et al. 2011, 2012, 2017; Ishiyama et al. 2004; Zhong et al. 2011). This has generated substantial public health concerns resulting in the release of statements from health regulatory agencies and clinical societies advocating that in the absence of direct evidence of harm with COVID-19, these medicines should not be discontinued (European Medicines Agency n.d.; European Society of Cardiology n.d.). However, inconsistencies in recommendations emerged with suggestions that users should either be monitored more closely (Fang, Karakiulakis, and Roth 2020) or that clinical trials investigating their withdrawal should be performed (Bauer and Massberg n.d.). Withholding these medicines, however, may result in worse cardiovascular outcomes with some studies reporting an increased risk of myocardial injury resulting from illness with COVID-19 (Shi et al. 2020).

Several studies have emerged examining this conundrum. While informative, they have either had smaller sample size, used heterogeneous study designs or had methodological limitations (Zhang et al. 2020; Dooley et al. n.d.). For example, all except two recent studies have so far directly compared the risk of COVID-19 with ACE or ARB use with an unexposed control population (de Abajo et al. 2020; Khera et al. 2020). This can result in non-comparable subjects, confounding by indication and the lack of a clear index date for when follow-up should start, all of which may induce bias.

Reliable evidence should also be replicable, generalizable and robust. To draw strong conclusions from observational studies, we must observe consistent findings produced from transparent, well-designed analyses across multiple populations and data capture processes to ensure that any associations are not due to systematic error or applicable only in narrow contexts. This study aimed to determine whether exposure to ACEs/ARBs is associated with an increased susceptibility to COVID-19 using a large multi-national federated active comparator cohort study facilitated by a common data model using the Observational Health Data Sciences and Informatics (OHDSI) (Hripcsak et al. 2015) network.

## Methods

### Study design

An international team of clinical, academic, government and industry stakeholders openly drafted the protocol for the International Covid-ACE Receptor Inhibition Utilization and Safety (ICARIUS) studies (https://github.com/ohdsi-studies/Covid19Icarius) and registered the protocol in the EU PAS register (EUPAS35296). Under our protocol, we conducted a systematic and comprehensive prevalent-user active comparator cohort study measuring the association between RAS use and susceptibility to COVID-19 among hypertensive patients using antihypertensive medication.

### Data sources

We identified patients in routinely-collected electronic health records (EHRs) and claims data from the United States (US) and Spain. All data sources had been mapped to the Observational Medical Outcomes Partnership (OMOP) Common Data Model (CDMv5) that the open-science OHDSI collaborative maintains (Hripcsak et al. 2015). Two particular benefits of this standardization are that contributing centers can participate in distributed network analyses without needing to share patient-level information and that we can ensure data provenance, while applying common analytical code across all data sources in a consistent manner. The data sources included:

- Columbia University Irving Medical Center data warehouse (CUIMC) EHRs covering approximately six million patients from the New York-Presbyterian Hospital/Columbia University Irving Medical Center in the US. CUIMC includes data on clinical diagnoses, prescriptions, laboratory tests, demographics, and COVID-19 tests and diagnosis;
- Information Systems for Research in Primary Care (SIDIAP) database, covering approximately 80% of the population of Catalonia, Spain, with approximately six million patients. SIDIAP contains data since 2006 from general practice EHRs linked to hospital admissions with information on diagnoses, prescriptions, laboratory tests, and lifestyle and sociodemographics and the central database of RT-PCR COVID-19 tests; and
- US Department of Veterans Affairs (VA) database, covering approximately 12 million patients from 170 medical centers across the US and including administrative, clinical, laboratory, and pharmacy data repositories that are linked using unique patient identifiers.

Each site obtained institutional review board approval or obtained a determination that the de-identified data were not human subjects research.

### Cohort eligibility, study period and follow-up

Each cohort consisted of adults aged 18 years or older who received at least one eligible outpatient prescription for an antihypertensive drug between 1^st^ November 2019 and 31^st^ January 2020. The index date was set as the date of the last prescription in this time window. We required patients to be observable in their data source for at least one year prior to the index date and have a recorded history of hypertension at any point prior to or including the index date (Figure 1). Cohort exit was the earliest of: the occurrence of an outcome; the end of exposure; death; loss or deregistration from the database; or date of last data collection.

**Figure 1.**
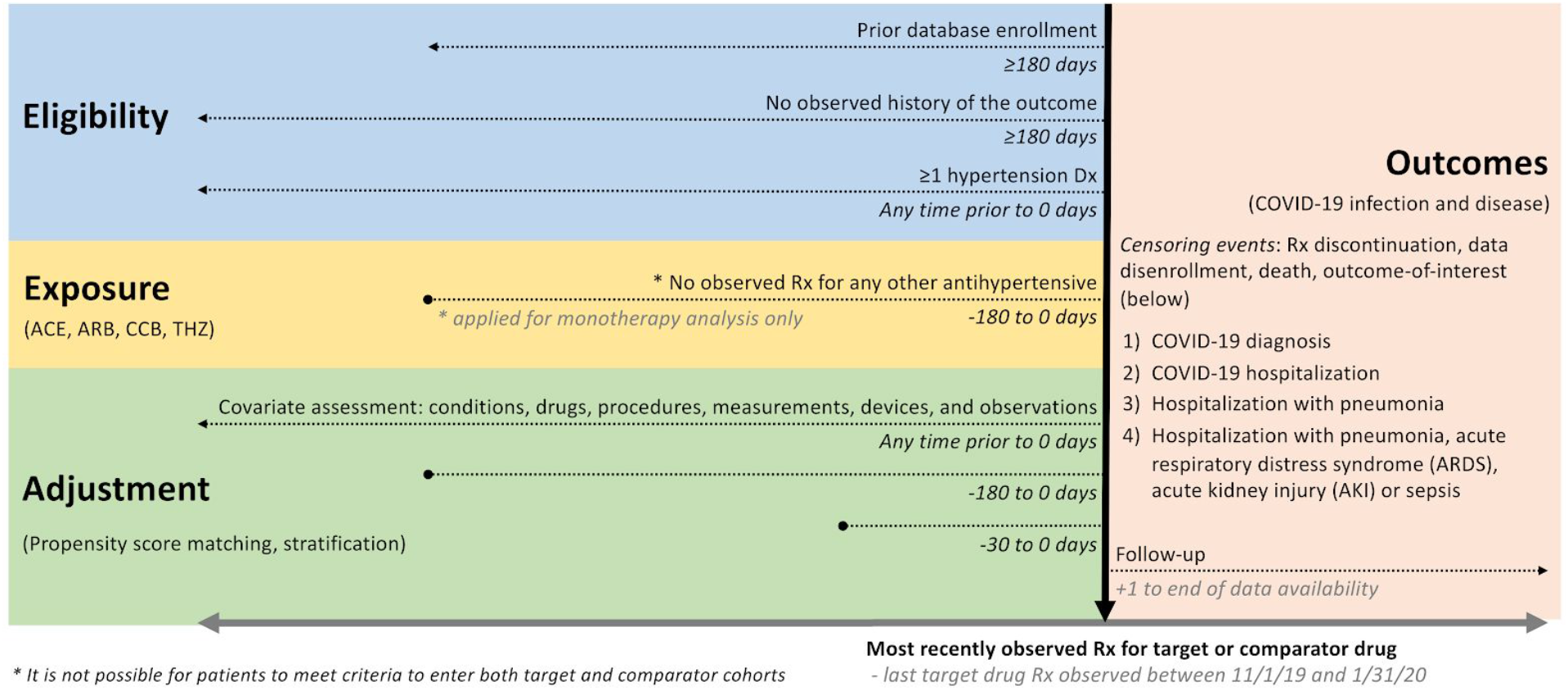
International Covid-ACE Receptor Inhibition Utilization and Safety (ICARIUS) susceptibility study design schematic. We highlight eligibility criteria, exposure definitions, adjustment strategies, index date specification (horizontal black arrow) and outcome definitions and time-at-risk. Exposure involves prescriptions to drugs with RxNorm ingredients that map to the first-line antihypertensive angiotensin converting enzyme inhibitor (ACE), angiotensin receptor blocker (ARB), dihydropyridine calcium channel blockers (CCB) and thiazide or thiazide-like diuretic (THZ) classes.

### Exposures

The exposures of interest were four first-line antihypertensive drug classes: ACEs, ARBs, dihydropyridine calcium channel blockers (CCBs) and thiazide or thiazide-like diuretics (THZs), defined separately as either 1) monotherapy or 2) monotherapy or combination therapy (simply called combination in this paper) users with other antihypertensives. Our primary comparison examined outpatient RAS blocker exposure (ACE or ARB) to CCBs or THZs exposure (included as active comparators). Further investigation compared class exposure to ACEs to exposure to ARBs separately, and individual classes to various active comparators, leading to ten different target-comparator pairings for monotherapy and combo-therapy each, as listed in the online supplement. For patients on monotherapy, we required the absence of any other antihypertensive treatment between –180 days and 0 days prior to the index date. We defined continuous drug exposures from the start of follow-up by grouping sequential prescriptions that have at most 30 days gap between prescriptions, and defined end of exposure as the end of the last prescription’s drug supply in such a sequence.

### Outcomes

We investigated four COVID-19 related outcomes: 1) COVID-19 diagnosis; 2) COVID-19 hospitalization; 3) hospitalization with pneumonia and; 4) hospitalization with pneumonia, acute respiratory distress syndrome (ARDS), acute kidney injury (AKI) or sepsis (PAAS). In brief, positive tests results or diagnostic codes defined COVID-19 status. The full details of the participant cohorts and outcomes used for development and validation can be found in the online protocol.

### Study size

We undertook this study using all patients meeting the eligibility criteria within each database. We therefore performed no *a priori* sample size calculation; instead, we provide a minimum detectable rate ratio (MDRR) for each target-comparator-outcome triplet across each data source.

### Statistical methods

To adjust for potential measured confounding and improve the balance between comparison cohorts, we built large-scale propensity score (PS) models (Rosenbaum and Rubin 1983) for each comparison and data source using a consistent data-driven process through regularized regression (Tian, Schuemie, and Suchard 2018). This process used a large set of predefined baseline patient characteristics, including age, gender, race (US data) and other demographics and prior conditions, drug exposures, procedures and health service utilization behaviors, to provide the most accurate prediction of treatment and balance patient cohorts across many characteristics. For computational efficiency, we excluded all features that occurred in fewer than 0.1% of patients within the target and comparator cohorts prior to PS model fitting.

In separate analyses, we stratified into 5 PS quintiles or variable-ratio matched patients by PS, and used Cox proportional hazards models to estimate hazard ratios (HRs) between alternative target and comparator treatments for the risk of each outcome in each data source. The regression conditioned on the PS strata/matching-unit with treatment allocation as the sole explanatory variable. We aggregated HR estimates across data sources to produce meta-analytic estimates using a random-effects meta-analysis (DerSimonian and Laird 1986). For both monotherapy and combination use of the ACE, ARB, CCB, THZ, ACE or ARB (ACE/ARB) and CCB or THZ (CCB/THZ) class groups (10 pairwise comparisons) to study four outcomes in three data sources (plus one meta-analysis) using two PS-adjustment approaches, we generated 2 x 10 x 4 x (3 + 1) x 2 = 1280 study effects.

Residual study bias from unmeasured and systematic sources often remains in observational studies even after controlling for measured confounding through PS-adjustment (Schuemie et al. 2014, 2016). For each study effect, we conducted negative control outcome experiments, where the null hypothesis of no effect is believed to be true, using 76 controls identified through a data-rich algorithm (Voss et al. 2017) and validated in a previous antihypertensive comparative study (Suchard et al. 2019). Using the empirical null distributions from these experiments, we calibrated each study effect HR estimate, its 95% confidence interval (CI) and the *p*-value to reject the null hypothesis of no differential effect (Schuemie et al. 2018). We declared a HR as significantly different from no effect when its calibrated *p* < 0.05 without correcting for multiple testing.

Blinded to the results, clinicians and epidemiologists evaluated study diagnostics for these treatment comparisons to assess if they were likely to yield unbiased estimates. The suite of diagnostics included (1) MDRR, (2) preference score (a transformation of the PS that adjusts for prevalence differences between populations) distributions to evaluate empirical equipoise (Walker et al. 2013) and population generalizability, (3) extensive patient characteristics to evaluate cohort balance before and after PS-adjustment, (4) negative control calibration plots to assess residual bias, and (5) Kaplan-Meier plots to examine HR proportionality assumptions. We defined target and comparator cohorts to stand in empirical equipoise if the majority of patients in both carry preference scores between 0.3 and 0.7 and to achieve sufficient balance if all after-adjustment baseline characteristics return absolute standardized mean differences (SMD) < 0.1 (Austin 2009).

### Study execution

We conducted this study using the open-source OHDSI CohortMethod R package (https://ohdsi.github.io/CohortMethod/) with large-scale analytics made possible through the Cyclops R package (Suchard et al. 2013). The pre-specified ICARIUS protocol and start-to-finish open and executable source code are available at: https://github.com/ohdsi-studies/Covid19Icarius. To promote transparency and facilitate sharing and exploration of the complete result set, an interactive web application (https://data.ohdsi.org/IcariusSusceptibility) serves up study diagnostics and results for all study effects.

## Results

### Population and incidence

A total of 363,785 hypertensive patients exposed to ACE/ARB monotherapy were compared to 248,915 CCB/THZ monotherapy users contributing 121,213 and 81,261 person years (pyrs) of follow-up respectively. The overall incidence of COVID-19 diagnosis among monotherapy was 5.6 vs 4.8 per 1,000 pyrs among ACE/ARB and CCB/THZ users respectively, although incidence rates varied by data source.

Corresponding patient cohort size and diagnosis incidence rates were: 268,711 and 5.6 per 1,000 pyrs for ACE (alone) monotherapy users and 92,485 and 5.1 per 1,000 pyrs for ARB (alone) monotherapy, when compared to CCB/THZ monotherapy users or each other. Cohorts for combination users (as monotherapy or in-combination) ranged as large as 711,799 for ACE/ARB users and 473,076 for CCB/THZ users.

The aggregated patient cohort size, follow-up duration, incidences of each COVID-19 related outcome and MDRR for each drug comparison and database are shown in Table 1. Supplementary Tables 1 - 4 provide further cohort size and outcome event information for all 10 pairwise cohort comparisons across all 4 outcomes.

**Table 1:**
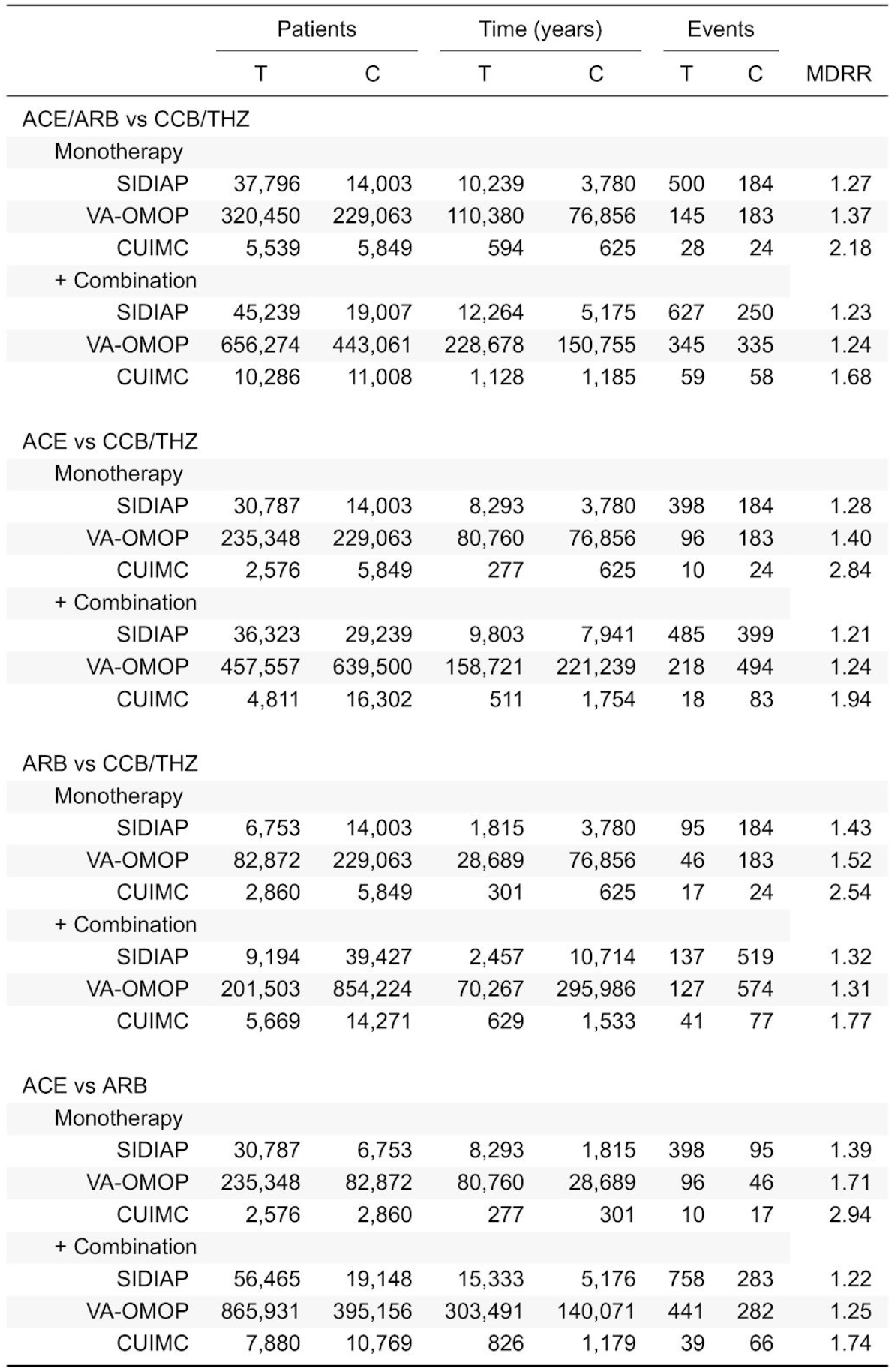
Populations and COVID-19 diagnoses for angiotensin converting enzyme (ACE) inhibitor, angiotensin receptor blocker (ARB), calcium channel block (CCB) and thiazide or thiazide-like diuretics (THZ) monotherapy and in-combination user cohorts. For each target (T) and comparator (C) cohort, we report population size, total exposure time, outcome events and minimally detectable risk ratio (MDRR).

|  | Patients |  | Time (years) |  | Events |  | MDRR |
| --- | --- | --- | --- | --- | --- | --- | --- |
|  | T | C | T | C | T | C |  |
| ACE/ARB vs CCB/THZ |  |  |  |  |  |  |  |
| Monotherapy |  |  |  |  |  |  |  |
| SIDIAP | 37,796 | 14,003 | 10,239 | 3,780 | 500 | 184 | 1.27 |
| VA-OMOP | 320,450 | 229,063 | 110,380 | 76,856 | 145 | 183 | 1.37 |
| CUIMC | 5,539 | 5,849 | 594 | 625 | 28 | 24 | 2.18 |
| + Combination |  |  |  |  |  |  |  |
| SIDIAP | 45,239 | 19,007 | 12,264 | 5,175 | 627 | 250 | 1.23 |
| VA-OMOP | 656,274 | 443,061 | 228,678 | 150,755 | 345 | 335 | 1.24 |
| CUIMC | 10,286 | 11,008 | 1,128 | 1,185 | 59 | 58 | 1.68 |
| ACE vs CCB/THZ |  |  |  |  |  |  |  |
| Monotherapy |  |  |  |  |  |  |  |
| SIDIAP | 30,787 | 14,003 | 8,293 | 3,780 | 398 | 184 | 1.28 |
| VA-OMOP | 235,348 | 229,063 | 80,760 | 76,856 | 96 | 183 | 1.40 |
| CUIMC | 2,576 | 5,849 | 277 | 625 | 10 | 24 | 2.84 |
| + Combination |  |  |  |  |  |  |  |
| SIDIAP | 36,323 | 29,239 | 9,803 | 7,941 | 485 | 399 | 1.21 |
| VA-OMOP | 457,557 | 639,500 | 158,721 | 221,239 | 218 | 494 | 1.24 |
| CUIMC | 4,811 | 16,302 | 511 | 1,754 | 18 | 83 | 1.94 |
| ARB vs CCB/THZ |  |  |  |  |  |  |  |
| Monotherapy |  |  |  |  |  |  |  |
| SIDIAP | 6,753 | 14,003 | 1,815 | 3,780 | 95 | 184 | 1.43 |
| VA-OMOP | 82,872 | 229,063 | 28,689 | 76,856 | 46 | 183 | 1.52 |
| CUIMC | 2,860 | 5,849 | 301 | 625 | 17 | 24 | 2.54 |
| + Combination |  |  |  |  |  |  |  |
| SIDIAP | 9,194 | 39,427 | 2,457 | 10,714 | 137 | 519 | 1.32 |
| VA-OMOP | 201,503 | 854,224 | 70,267 | 295,986 | 127 | 574 | 1.31 |
| CUIMC | 5,669 | 14,271 | 629 | 1,533 | 41 | 77 | 1.77 |
| ACE vs ARB |  |  |  |  |  |  |  |
| Monotherapy |  |  |  |  |  |  |  |
| SIDIAP | 30,787 | 6,753 | 8,293 | 1,815 | 398 | 95 | 1.39 |
| VA-OMOP | 235,348 | 82,872 | 80,760 | 28,689 | 96 | 46 | 1.71 |
| CUIMC | 2,576 | 2,860 | 277 | 301 | 10 | 17 | 2.94 |
| + Combination |  |  |  |  |  |  |  |
| SIDIAP | 56,465 | 19,148 | 15,333 | 5,176 | 758 | 283 | 1.22 |
| VA-OMOP | 865,931 | 395,156 | 303,491 | 140,071 | 441 | 282 | 1.25 |
| CUIMC | 7,880 | 10,769 | 826 | 1,179 | 39 | 66 | 1.74 |

### Characteristics of patients

Baseline characteristics of ACE/ARB monotherapy users compared to CCB/THZ monotherapy users, before and after PS stratification, are shown in Table 2. There were baseline differences in sex, hyperlipidaemia, diabetes, renal impairment, heart failure, heart disease, atrial fibrillation, drugs for diabetes, lipid modifying agents, antithrombotics, antacids, opioids, and race that varied by data source. Further information on the population characteristics for each cohort comparison and design evaluated for each data source are shown in Supplementary Tables 5 - 64, one for each of the 2 ⨯ 10⨯ 3 = 60 comparisons across data sources.

**Table 2:**
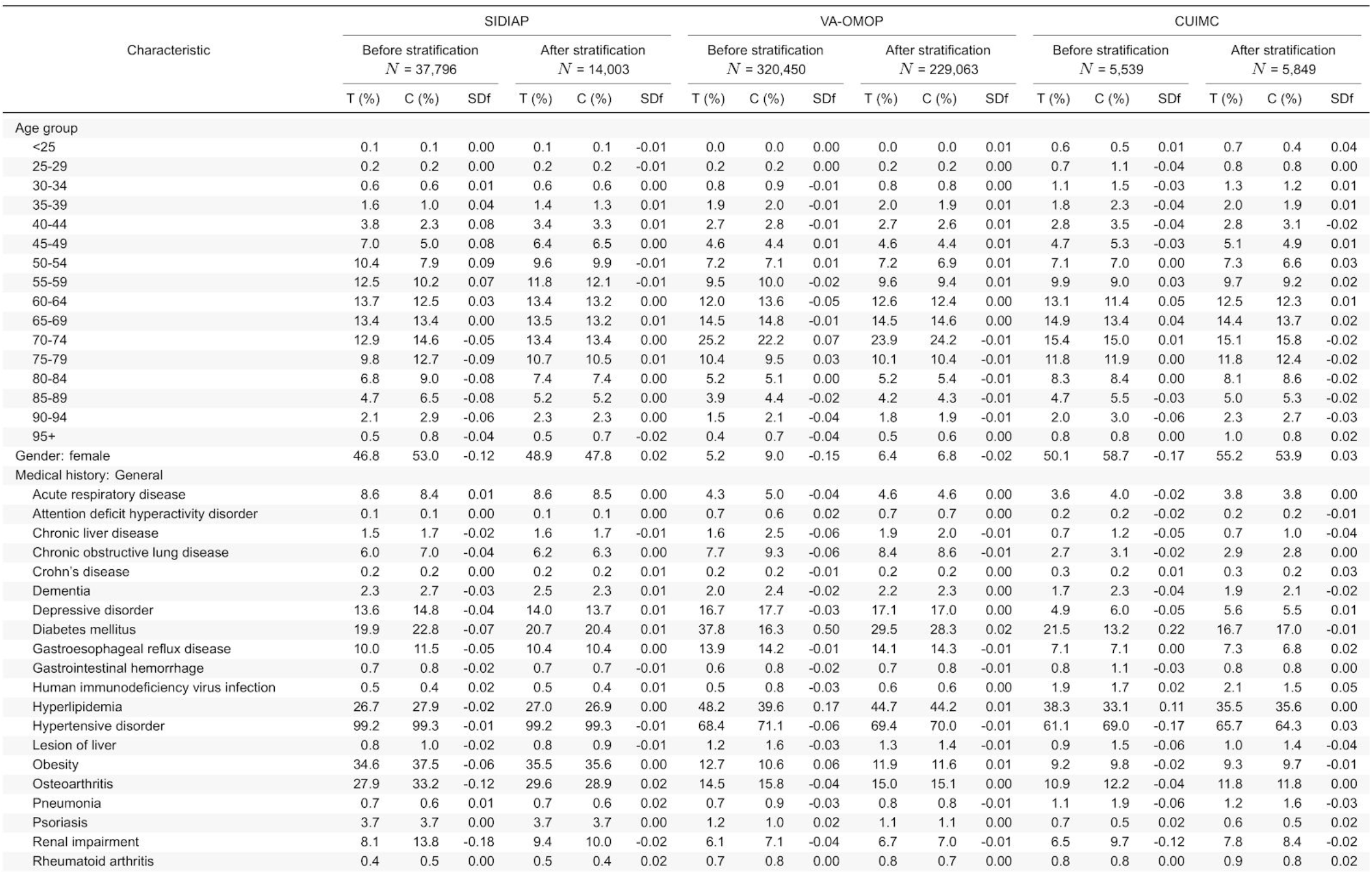
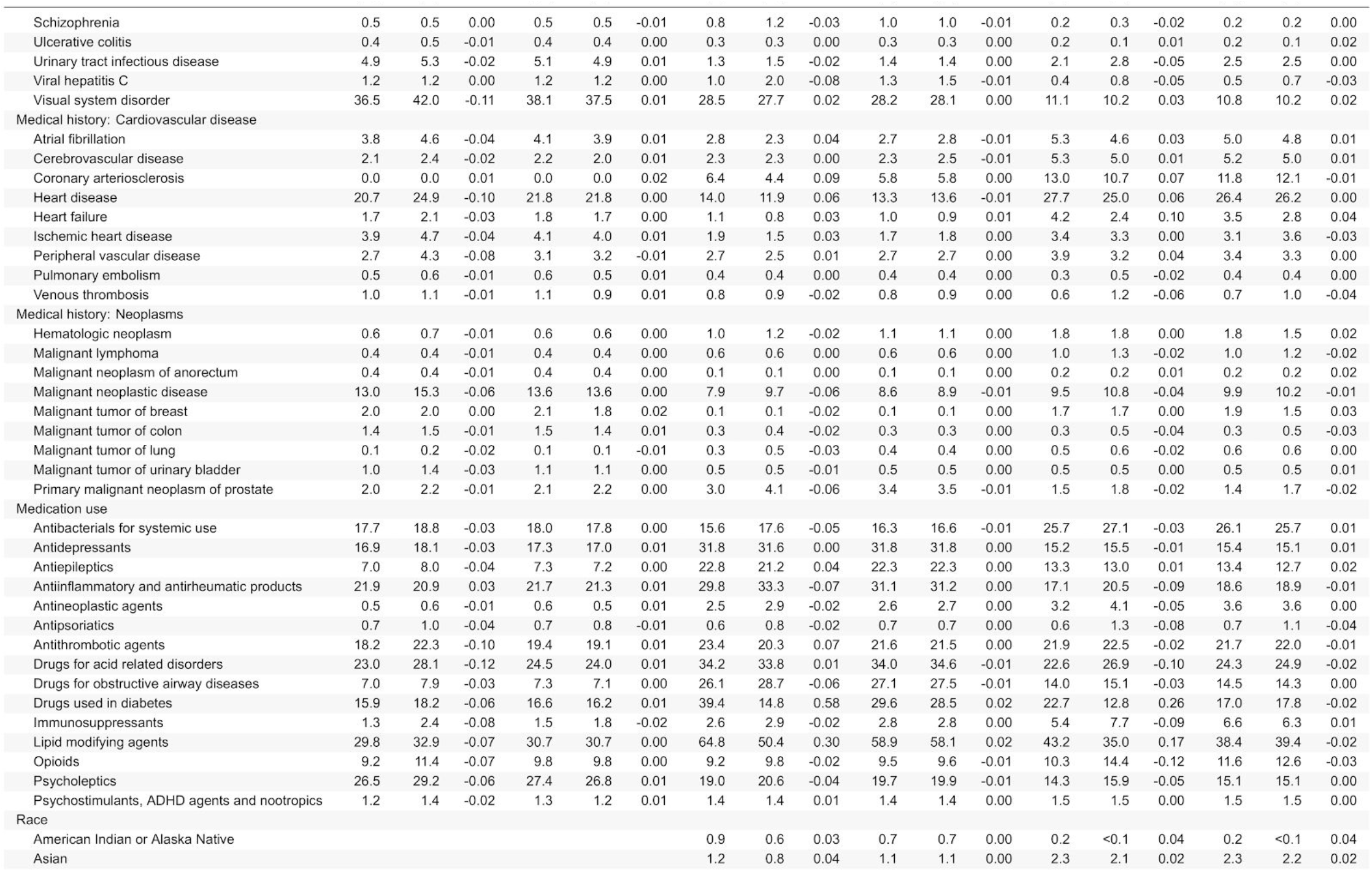
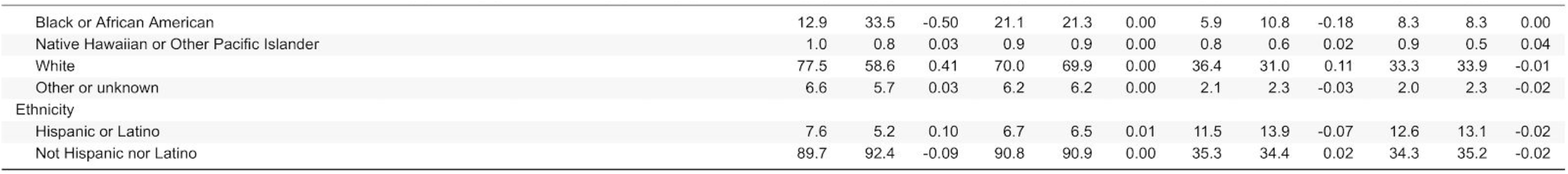
Baseline patient characteristics for ACE/ARB (T) and CCB/THZ (C) monotherapy prevalent-use in the SIDIAP, VA-OMOP and CUIMC data sources. We report the proportion of use satisfying selected based-line characteristics and the standardized difference of population proportions (SDf) before and after stratification. Less extreme SDf through stratification suggest improved balance between patient cohorts through propensity score adjustment.

### PS model adjustment and cohort balance

The number of baseline patient characteristics available differed across comparison-cohorts and data sources, ranging from 6,447 to 9,860 in SIDIAP, 10,638 to 12,425 in VA-OMOP and 11,776 to 21,440 in CUIMC. After large-scale PS construction and then stratification or matching, SMDs for all baseline characteristics were <0.1 in SIDIAP and VA-OMOP for each drug comparison, apart from the comparison between combination users of ARBs and CCBs/THZs in VA-OMOP. SMDs for all baseline characteristics before and after PS adjustment for ACE/ARB monotherapy users compared to CCB/THZ monotherapy users for all data sources are plotted in Figure 2. In CUIMC, all but one drug comparison (ACE vs ARB monotherapy) with PS stratification showed residual cohort imbalances with a SMD ≥0.1, which involved baseline characteristics related to pregnancy, renal transplantation, and heart failure and use of sacubitril. However, these cohort comparisons all passed study diagnostics for the PS matching design. Supplementary Figures 1 - 60 demonstrate study diagnostics for all comparisons and include negative control effect estimate distributions.

**Figure 2.**
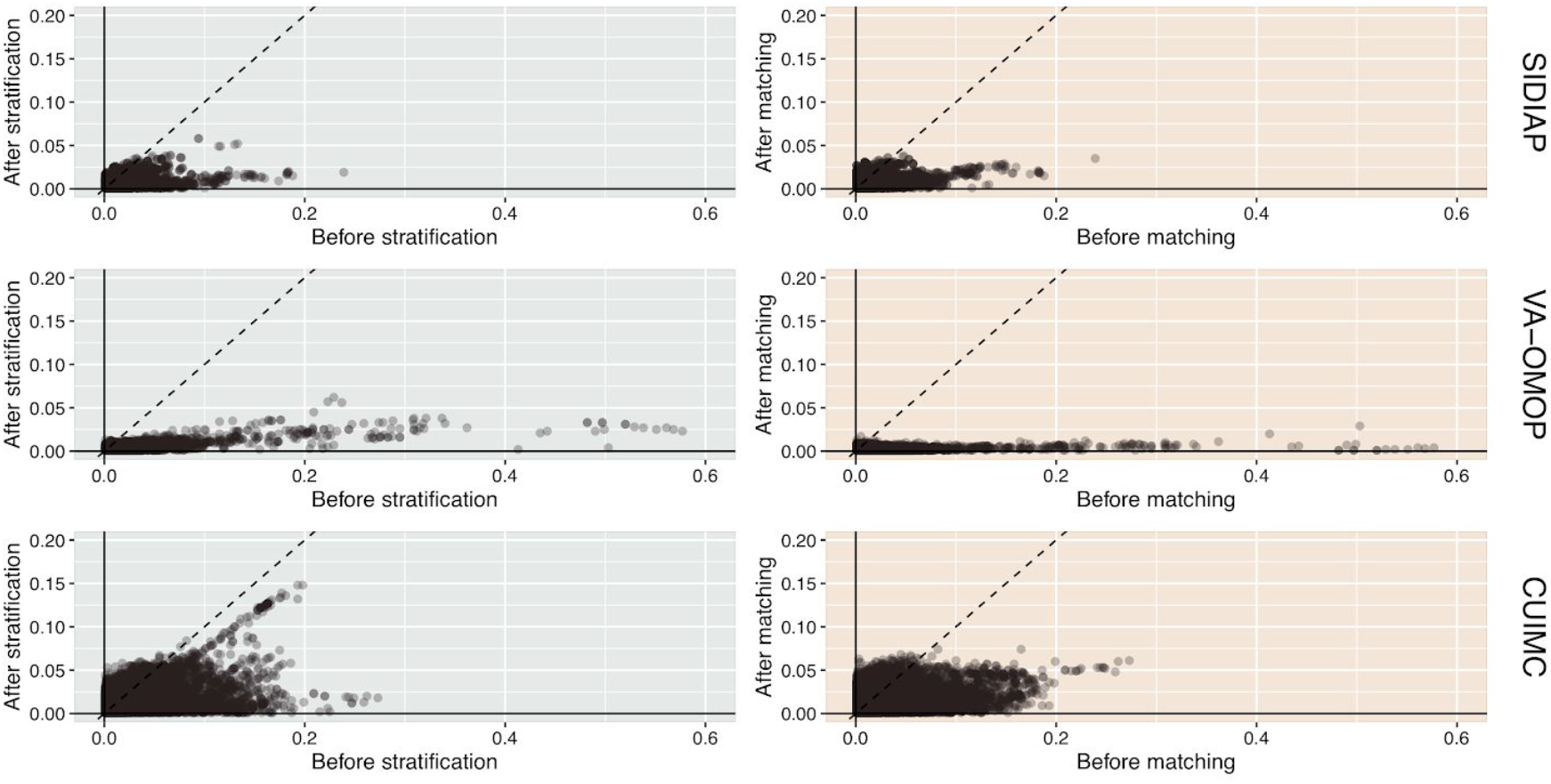
Cohort balance diagnostics comparing ACE/ARB and CCB/THZ monotherapy prevalent-use for the risk COVID-19 diagnosis. We plotted the absolute standardized difference of population proportions of all available patient characteristics (6,571 in SIDIAP, 11,183 in VA-OMOP, and 18,291 in CUIMC) before and after propensity score stratification or matching across data sources. Using stratification, CUIMC fails study diagnostics for this comparison as the absolute standardized difference is not consistently <0.1.

### Risk of COVID-19 diagnosis

Calibrated HRs for the relative risk of incident COVID-19 diagnosis are presented in Table 3 and Figure 3 for PS stratified and PS matched analyses. In SIDIAP, the risk of of COVID-19 diagnosis was not significantly different among ACE/ARB users compared to CCB/THZ use with PS stratification (HR 1.02, 95%CI 0.86-1.21 for monotherapy and HR 1.06, 95%CI 0.92-1.24 with combination use). Corresponding hazard ratios in VA-OMOP were HR 0.91 (95%CI 0.71-1.17) and HR 0.98 (95%CI 0.81-1.18). PS stratification did not pass study diagnostics in CUIMC and is therefore grayed out in the table to caution against interpretation. The corresponding HRs for CUIMC using PS matching were HR 0.67 (95%CI 0.20-2.20) and HR 2.36 (95%CI 0.98-5.68) respectively. Meta-analytic HRs following PS stratification for ACE/ARB use were 0.98 (95%CI 0.84 - 1.14) for monotherapy, and 1.01 (95%CI 0.90 - 1.15) with combination use.

**Table 3.**
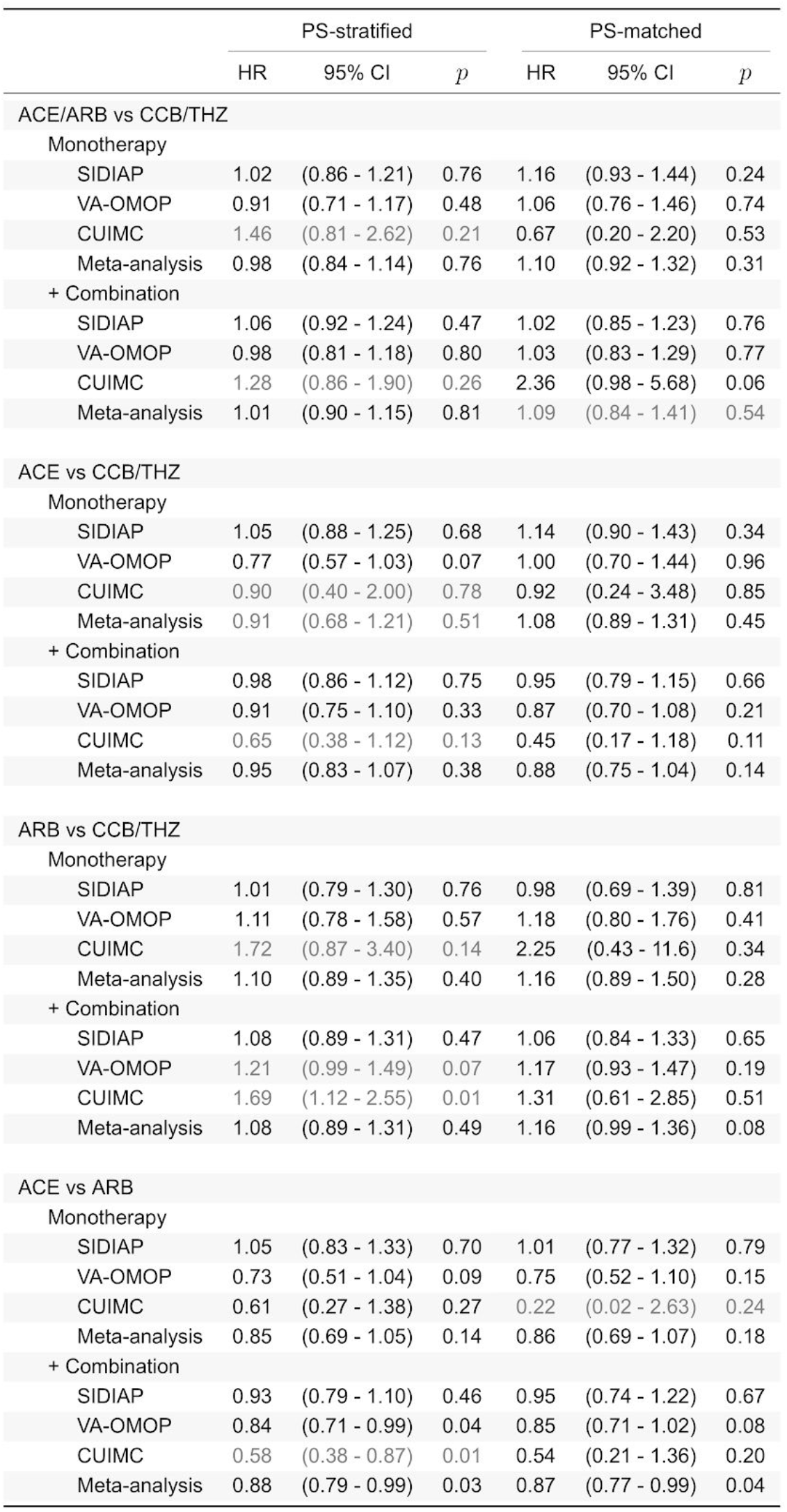
Relative risk of COVID-19 diagnosis for ACE inhibitor, ARB, CCB and THZ prevalent-use. We report calibrated hazard ratios (HRs) and their 95% confidence intervals (CIs) and calibrated p-value (P) for both monotherapy and in-combination cohort definitions, with propensity score (PS) stratification or matching and across data sources. Grayed out entries do not pass study diagnostics

|  | PS-stratified |  |  | PS-matched |  |  |
| --- | --- | --- | --- | --- | --- | --- |
|  | HR | 95% CI | p | HR | 95% CI | p |
| ACE/ARB vs CCB/THZ |  |  |  |  |  |  |
| Monotherapy |  |  |  |  |  |  |
| SIDIAP | 1.02 | (0.86 - 1.21) | 0.76 | 1.16 | (0.93 - 1.44) | 0.24 |
| VA-OMOP | 0.91 | (0.71 - 1.17) | 0.48 | 1.06 | (0.76 - 1.46) | 0.74 |
| CUIMC | 1.46 | (0.81 - 2.62) | 0.21 | 0.67 | (0.20 - 2.20) | 0.53 |
| Meta-analysis | 0.98 | (0.84 - 1.14) | 0.76 | 1.10 | (0.92 - 1.32) | 0.31 |
| + Combination |  |  |  |  |  |  |
| SIDIAP | 1.06 | (0.92 - 1.24) | 0.47 | 1.02 | (0.85 - 1.23) | 0.76 |
| VA-OMOP | 0.98 | (0.81 - 1.18) | 0.80 | 1.03 | (0.83 - 1.29) | 0.77 |
| CUIMC | 1.28 | (0.86 - 1.90) | 0.26 | 2.36 | (0.98 - 5.68) | 0.06 |
| Meta-analysis | 1.01 | (0.90 - 1.15) | 0.81 | 1.09 | (0.84 - 1.41) | 0.54 |
| ACE vs CCB/THZ |  |  |  |  |  |  |
| Monotherapy |  |  |  |  |  |  |
| SIDIAP | 1.05 | (0.88 - 1.25) | 0.68 | 1.14 | (0.90 - 1.43) | 0.34 |
| VA-OMOP | 0.77 | (0.57 - 1.03) | 0.07 | 1.00 | (0.70 - 1.44) | 0.96 |
| CUIMC | 0.90 | (0.40 - 2.00) | 0.78 | 0.92 | (0.24 - 3.48) | 0.85 |
| Meta-analysis | 0.91 | (0.68 - 1.21) | 0.51 | 1.08 | (0.89 - 1.31) | 0.45 |
| + Combination |  |  |  |  |  |  |
| SIDIAP | 0.98 | (0.86 - 1.12) | 0.75 | 0.95 | (0.79 - 1.15) | 0.66 |
| VA-OMOP | 0.91 | (0.75 - 1.10) | 0.33 | 0.87 | (0.70 - 1.08) | 0.21 |
| CUIMC | 0.65 | (0.38 - 1.12) | 0.13 | 0.45 | (0.17 - 1.18) | 0.11 |
| Meta-analysis | 0.95 | (0.83 - 1.07) | 0.38 | 0.88 | (0.75 - 1.04) | 0.14 |
| ARB vs CCB/THZ |  |  |  |  |  |  |
| Monotherapy |  |  |  |  |  |  |
| SIDIAP | 1.01 | (0.79 - 1.30) | 0.76 | 0.98 | (0.69 - 1.39) | 0.81 |
| VA-OMOP | 1.11 | (0.78 - 1.58) | 0.57 | 1.18 | (0.80 - 1.76) | 0.41 |
| CUIMC | 1.72 | (0.87 - 3.40) | 0.14 | 2.25 | (0.43 - 11.6) | 0.34 |
| Meta-analysis | 1.10 | (0.89 - 1.35) | 0.40 | 1.16 | (0.89 - 1.50) | 0.28 |
| + Combination |  |  |  |  |  |  |
| SIDIAP | 1.08 | (0.89 - 1.31) | 0.47 | 1.06 | (0.84 - 1.33) | 0.65 |
| VA-OMOP | 1.21 | (0.99 - 1.49) | 0.07 | 1.17 | (0.93 - 1.47) | 0.19 |
| CUIMC | 1.69 | (1.12 - 2.55) | 0.01 | 1.31 | (0.61 - 2.85) | 0.51 |
| Meta-analysis | 1.08 | (0.89 - 1.31) | 0.49 | 1.16 | (0.99 - 1.36) | 0.08 |
| ACE vs ARB |  |  |  |  |  |  |
| Monotherapy |  |  |  |  |  |  |
| SIDIAP | 1.05 | (0.83 - 1.33) | 0.70 | 1.01 | (0.77 - 1.32) | 0.79 |
| VA-OMOP | 0.73 | (0.51 - 1.04) | 0.09 | 0.75 | (0.52 - 1.10) | 0.15 |
| CUIMC | 0.61 | (0.27 - 1.38) | 0.27 | 0.22 | (0.02 - 2.63) | 0.24 |
| Meta-analysis | 0.85 | (0.69 - 1.05) | 0.14 | 0.86 | (0.69 - 1.07) | 0.18 |
| + Combination |  |  |  |  |  |  |
| SIDIAP | 0.93 | (0.79 - 1.10) | 0.46 | 0.95 | (0.74 - 1.22) | 0.67 |
| VA-OMOP | 0.84 | (0.71 - 0.99) | 0.04 | 0.85 | (0.71 - 1.02) | 0.08 |
| CUIMC | 0.58 | (0.38 - 0.87) | 0.01 | 0.54 | (0.21 - 1.36) | 0.20 |
| Meta-analysis | 0.88 | (0.79 - 0.99) | 0.03 | 0.87 | (0.77 - 0.99) | 0.04 |

**Figure 3.**
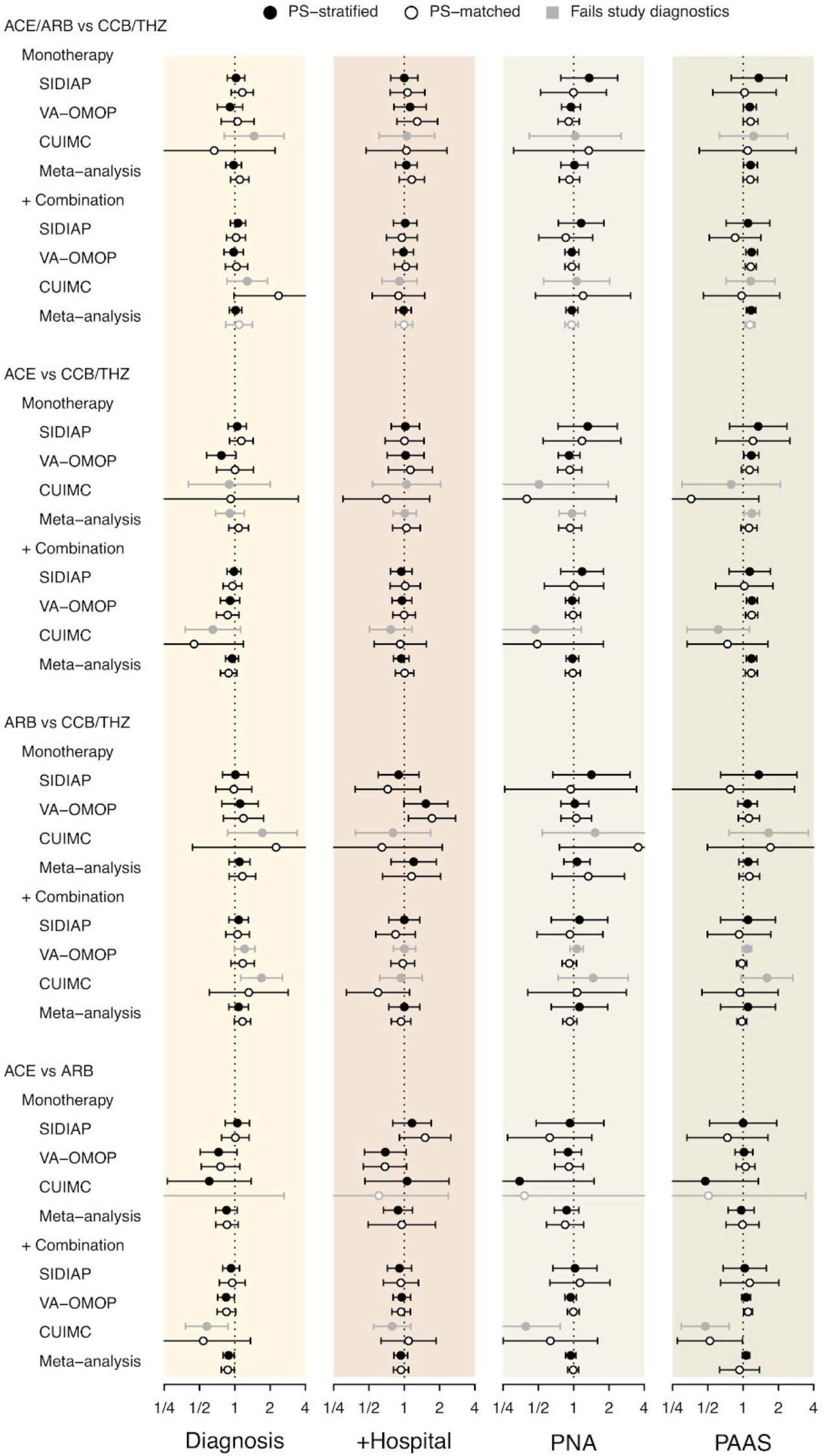
Relative risk of COVID-19-related outcomes for ACE inhibitor, ARB, CCB and THZ prevalent-use across data sources. Outcomes are COVID-19 diagnosis, COVID-19 hospitalization (+Hospital), patients hospitalized with pneumonia (PNA) and patients hospitalized with pneumonia, acute respiratory distress syndrome, acute kidney injury or sepsis (PAAS). We plot calibrated hazard ratios (circles) and their 95% confidence intervals labeled by propensity score (PS) adjustment method (black/white). Grayed out entries do not pass study diagnostics.

When comparing ACE and ARB use separately to CCB/THZ use, we observed no significant difference with COVID-19 diagnosis for comparisons passing study diagnostics (Table 3). For ACE use, meta-analytic HRs for monotherapy were 0.91 (95%CI 0.68 - 1.21), but with heterogeneity > 40%, and 0.95 (95%CI 0.83 - 1.08) for combination use. For ARB use, meta-analytic HRs for monotherapy were 1.10 (95%CI 0.89 - 1.35) for PS stratification, and 1.08 (95%CI 0.89 - 1.31) with combination use.

When comparing ACE use directly to ARB use, no significant difference in the risk of COVID-19 diagnosis was observed in individual databases, apart from combination use in VA-OMOP (HR 0.84, 95%CI 0.71 - 0.99). Meta-analytic HRs for monotherapy were 0.85 (95%CI 0.69 - 1.05) for PS stratification and 0.88 (95%CI 0.79 - 0.99) for combination use. PS matching, where comparisons from CUIMC passed all PS diagnostics, produced similar results (Table 3).

### Risk of COVID-19 hospitalization, pneumonia and PAAS

Calibrated HRs for the risk of COVID-19 hospitalization are presented in Figure 3. We observed no significant association between incident COVID-19 hospitalization for the comparison with ACE/ARB use, evaluated either together or separately, compared to CCB/THZ use. For ACE use compared to ARB use using PS stratification, meta-analytic HRs for monotherapy were 0.88 (95%CI 0.66 - 1.17) and 0.93 (95%CI 0.82 - 1.07) with combination use.

No significant associations with the risk of pneumonia were observed with any drug comparison that satisfied study diagnostics. No significant associations with the risk of PAAS were observed with any drug comparison that satisfied study diagnostics in SIDIAP and CUIMC. In VA-OMOP no significant difference was observed in comparisons between ARB versus CCB/THZ use or ACE versus ARB use, while small significant associations were observed with ACE versus CCB/THZ (Supplementary Table 192).

## Discussion

### Summary of results

In this multicenter cohort study following more than 1.1 million hypertensive patients from the US and Spain, we observed no clear increased risk of COVID-19 diagnosis, hospitalization, or subsequent complications associated with the outpatient prevalent ACE or ARB use. The clinical implication of this study is that patients should not halt their ACE or ARB therapy despite previously posited mechanisms of increased COVID-19 risk.

We observed one nominally significant meta-analysis difference, that use of ACE combination therapy had a lower risk of COVID-19 diagnosis when compared with ARB combination therapy use. There was, however, no corresponding difference detected in hospitalization or complications. Therefore, the observed association may be due to chance or residual bias. Even if true, there is only a 12% difference and therefore favoring ACE over ARB for mitigating COVID-19 is not strongly supported by our result.

### Comparison with other literature

Four studies assessing the risk of Covid-19 infection among ACE/ARB use have been published originating from Italy, Spain and the US (de Abajo et al. 2020; Gnavi et al. 2020; Mancia et al. 2020; Mehra et al. 2020a; Reynolds et al. 2020). After adjustment for the higher prevalence of cardiovascular conditions in COVID-19 patients, ACE and ARB use was not associated with an increased risk of COVID-19 diagnosis. These case-control studies included only a limited number of covariates for model adjustment. Only two studies have compared the risk of COVID-19 susceptibility in ACE/ARB use with an active comparator (de Abajo et al. 2020; Khera et al. 2020a). In this context, comparing patients with similarly-indicated treatments is critical for reducing the risk of bias resulting from confounding by indication (e.g. hypertension), where the absence of treatment indicates either too mild disease to warrant pharmacological treatment (e.g. mild hypertension under control with lifestyle and diet changes), the presence of contraindications, or extreme frailty precluding the use of preventative medicines (e.g. at end-of-life) (Crump et al. 2009; Glynn et al. 2001; Petersen et al. 2012; Yoshida, Solomon, and Kim 2015). Indeed de Abajo et al. clearly demonstrates that compared to other antihypertensive use, non-use was associated with a significantly reduced risk of COVID-19 hospitalization with estimated odds ratios for severe and less severe cases of 0·48 (0·34–0·69) and 0·57 (0·43–0·75) respectively (de Abajo et al. 2020).

While no previous study has directly compared the risk of COVID-19 diagnosis between ACE and ARB use, several studies have reported lower point estimates associated with ACEs than for ARBs. These odds ratios range from 0.61 to 0.92 for ACEs and 0.89 to 1.10 for ARB (Mehta et al. 2020; Khera et al. 2020b; Gnavi et al. 2020; de Abajo et al. 2020). However, not all observational studies have suggested a differential effect between ACE and ARB use (Mancia et al. 2020).

ACEs and ARBs have different effects on angiotensin II, the primary substrate of ACE2, required by SARS-CoV-2 to enter human cells (Vaduganathan et al. 2020). Animal models have suggested that while ACEs increase ACE2 gene expression, they do not alter ACE2 activity, unlike ARBs, providing a potential mechanism for why differential effects might occur (Ferrario et al. 2005); (Rice et al. 2004). However, recent studies in humans have identified no difference in ACE2 levels following exposure to ACE or ARB use (Emilsson et al. 2020); (Gill et al. 2020). Therefore, our findings could also be explained by residual confounding, as suggested by recent comparisons of the incidence of *Staphylococcus aureus* infection and other outcomes between ACE and ARB use which suggest that ARB is not a perfect comparator for ACE, although no large scale PS-adjustment was used (Bidulka et al. 2020).

Furthermore, one study has reported an increased risk of COVID-19 hospitalization and ICU admission associated with use of ACE and ARB (Mehta et al. 2020). While we did not observe a consistent increased risk of hospitalization, we did observe an increased risk of PAAS largely driven by ACE use compared to CCB/THZ use. This may be related to the higher incidence of AKI associated with ACE use as no increased risk was observed for pneumonia, and AKI would be significantly more frequent than ARDS or sepsis.

### Strengths and limitations

We used an open science community approach to design a set of analyses consistently applied across a network of observational databases to generate results that can be directly compared and interpreted in aggregate. These analyses employed active comparators to reduce confounding by indication and is the first to apply large-scale propensity adjustment with full diagnostics and a large set of negative control experiments. We published the study protocol ahead of time and kept results blinded when assessing PS diagnostics helping to address concerns about reproducibility, robustness and transparency that have recently emerged (Rubin 2020; Mehra et al. 2020). We examined outpatient prevalent antihypertensive use because a new-user design in the context of COVID-19 that has widely affected the provision of routine care is infeasible. Therefore mediators on the causal pathway between exposure and outcome may be included in the adjustment. This may not necessarily result in bias in this setting however, as COVID-19 is a new illness and will not have affected the decision to initiate one drug over another and no depletion of susceptibles will have occurred. Similarly, biological mechanisms relating to ACE2 expression may require chronic exposure, hindering a new-user design. Prior treatment remains highly correlated with many baseline features that our large-scale propensity model considers when balancing patients and can provide some protection against this potential bias.

Further, we defined COVID-19 diagnosis through the presence of diagnostic codes or test positive results that will underestimate the number of true COVID-19 cases, the extent of which will vary by site due to differences in testing strategies. To address this potential limitation, we included a hospitalization-based COVID-19 outcome and observed similar results. Finally, we defined drug exposures by recorded prescriptions and we have no information on adherence. While we have used a rigorous approach (Suchard et al. 2019) to observational research, residual confounding is still possible.

### Clinical implications and conclusions

Our findings stand in agreement with regulatory and clinical society advice that ACE and ARB therapy should be continued in light of COVID-19. Further, the marginal difference between ACEs and ARBs does not warrant class switching to reduce COVID-19 susceptibility.

## Data Availability

This study was conducted as a distributed database network analysis. To protect patient privacy, all patient-level data were maintained securely behind institutional firewalls. Analysis code was downloaded and executed by each participating data partner, which generated only aggregate summary statistics (cohort counts, model coefficients) which were then centralized and synthesized in preparation of this manuscript. All aggregate summary statistics produced have been made publicly available at:

https://data.ohdsi.org/IcariusSusceptibility/

https://github.com/ohdsi-studies/Covid19Icarius

## Acknowledgements

We acknowledge the tremendous work and dedication of the 350 participants from 30 nations in the March 2020 OHDSI COVID-19 Virtual Study-a-thon (https://www.ohdsi.org/covid-19-updates/), without whom this study could not have been realized.

## Ethical Approval

All data partners received IRB approval or waiver in accordance with their institutional governance guidelines. Use of the SIDIAP data source was approved by the Clinical Research Ethics Committee of IDIAPJGol (project code: 20/070-PCV). Use of the VA-OMOP data source was reviewed by the Department of Veterans Affairs Central Institutional Review Board (IRB) and was determined to meet the criteria for exemption under Exemption Category 4(3) and approved the request for Waiver of HIPAA Authorization. Use of the CUIMC data source was approved by the Columbia University Institutional Review Board as an OHDSI network study (IRB# AAAO7805).

## Supplementary Material

Available at: https://github.com/ohdsi-studies/Covid19Icarius/blob/master/Documents/SusceptibilitySupplement.pdf

